# Development of Integrated Physician and Patient Tools to Improve Management of COPD: The ACCEPT Decision Intervention

**DOI:** 10.64898/2026.11.24.26368807

**Authors:** Rebecca K Metcalfe, Alison M Hoens, Ramona Sharma, Jim Johnson, Samir Gupta, Laura Struik, Joan L Bottorff, Mehrshad Mokhtaran, Mohsen Sadatsafavi, Paloma Burns, Don D Sin, Nick Bansback, the IMPACT study

**Affiliations:** Centre for Advancing Health Outcomes, Providence Health Research Institute, Vancouver, Canada; Department of Physical Therapy, University of British Columbia, Vancouver, Canada; Department of Social Work, University of British Columbia, Kelowna, Canada; Faculty of Pharmaceutical Sciences, University of British Columbia, Vancouver, Canada; Li Ka Shing Knowledge Institute, St. Michael’s Hospital, Toronto, Canada; Department of Medicine, University of Toronto, Toronto, Canada; School of Nursing, University of British Columbia, Kelowna, Canada; PROOF Centre of Excellence, University of British Columbia, Vancouver, Canada; Centre for Heart Lung Innovation, St. Paul’s Hospital, Vancouver, Canada; School of Population and Public Health, University of British Columbia, Vancouver, Canada

**Keywords:** Chronic obstructive pulmonary disease, prediction models, decision support tool, patient decision tool, user-centered design

## Abstract

**Background:** Chronic obstructive pulmonary disease (COPD) is a leading cause of morbidity and mortality, with COPD exacerbations being a leading cause of hospitalizations in Canada and many other countries. Preventive pharmacotherapy for reducing exacerbation risk is an important aspect of disease management in contemporary guidelines. However, real-world application is hindered by inappropriate prescribing and low patient adherence. This study focuses on developing the ACCEPT Decision Intervention (ADI), a tool designed to enhance both physician prescribing and patient adherence in line with clinical guidelines.

**Methods:** The ADI, which incorporates both physician and patient components, was developed using the Knowledge to Action framework. The physician component integrates the Acute COPD Exacerbation Prediction Tool (ACCEPT), a risk scoring tool for exacerbations, into the electronic medical record, offering tailored treatment recommendations based on clinical outcomes. The patient component provides a personalized educational handout addressing medication information, inhaler techniques, and self-management strategies. Development was informed by a series of semi-structured interviews and surveys with pulmonologists and patients, with iterative prototyping based on feedback.

**Results:** Interviews revealed barriers such as physicians’ difficulty in risk assessment and patients’ misconceptions about medication, which were addressed in the ADI design. The physician tool simplifies guideline adherence by automating risk prediction, while the patient handout offers customized, accessible information to improve treatment adherence. Feedback from pulmonologists highlighted the importance of minimizing workflow disruption, leading to streamlined tool design. Cognitive interviews with patients resulted in revisions to enhance clarity and engagement, such as incorporating more visuals and simplified language.

**Conclusion:** The ADI is a comprehensive, user-centered decision support tool aimed at improving COPD management by communicating exacerbation risk and aligning physician prescribing with guidelines and empowering patients to adhere to treatment. By integrating multivariable risk scoring and behaviour change strategies, this intervention has the potential to reduce exacerbation rates and hospitalizations. Ongoing evaluation in the IMPACT trial will assess its clinical effectiveness and implementation feasibility.

## 1. Background

Chronic obstructive pulmonary disease (COPD) is a chronic airway disease whose natural history is characterized by recurrent episodes of symptom worsening, referred to exacerbations or lung attacks.^1,2^ Preventive pharmacotherapy for exacerbations is a cornerstone of contemporary management of COPD. Contemporary guidelines unanimously emphasize preventive pharmacotherapy based on systematic reviews demonstrating that appropriate treatment decreases patient risk of hospitalization and death.^3,4^ However, realizing these benefits in a ‘real-world’ context requires physicians to prescribe appropriate medications and patients to take medications as prescribed. In reality, guideline adherence is poor.^5,6^ Among prescribers, this poor adherence includes both under-and over-prescription. Recent analyses demonstrated that less than 1% of patients with COPD receiving inhaled corticosteroids met guideline criteria for this medication class,^7^ and only half were adherent to treatment.^8^ Although COPD is considered an “ambulatory-sensitive” condition. given these gaps in pharmacotherapy, COPD remains a leading cause of hospitalizations in North America.

The causes of inappropriate prescribing and low medication adherence differ and are complex. While there are multiple reasons physicians may not prescribe guideline-recommended treatment, one reason may be difficulty with accurately identifying a patient’s risk.^9^ Current clinical guidelines^3,4^ outlining optimal treatment use exacerbation history to identify patients at a relatively higher risk of future exacerbations, without quantifying that future risk based on other key variables. This might result in physician’s overriding guideline recommendations given their own judgment of future exacerbation risk being different from what exacerbation history alone might indicate. As well, medication adherence is known to be strongly influenced by patient beliefs about medications and engagement in treatment decision making,^10–12^ particularly among patients with low health literacy.^13^ Further, evidence shows that behaviour change approaches can support patients to meaningfully improve inhalation technique.^14^ Given this, quantifying and communicating future exacerbation risk has the potential to improve provider adherence to guideline-recommended therapies, and, if coupled with evidence-based behaviour change techniques, patient adherence to pharmacotherapy.

There are several multivariable risk scoring tools for COPD exacerbations.^15–17^ Computer-based decision support tools that can query electronic patient charts present an opportunity to overcome this barrier by automating multivariable risk predictions. Previous evidence has found that computer-based decision support tools can improve concordance between guidelines and physician behaviour.^18,19^ However, compared to exacerbation history alone, such tools require more effort to collect all predictors.

Addressing patient beliefs and priorities has been found to improve medication adherence in various settings. A review reported that clinician conversation and tailored patient materials, as well as ongoing support, were key components of effective outpatient self-management of COPD.^20^ Likewise, short decision tools have been shown to successfully support shared decision making during clinical encounters,^21^ and compared to usual care, shared decision making has been shown to improve both therapeutic adherence and disease outcomes in airways disease.^22^

The objective of this study was to develop the ACCEPT Decision Intervention (ADI), an integrated physician and patient decision tool to improve the appropriateness of physician prescribing as assessed by clinical guidelines, patient engagement in treatment decision making, and consequently, patient adherence to treatment.^23^ This decision support intervention was designed around a clinical prediction model called ACCEPT (<u>Ac</u>ute <u>C</u>OPD <u>E</u>xacerbation <u>P</u>rediction <u>T</u>ool), which uses up to 13 clinical variables (i.e., history of exacerbations, age, sex, body-mass index, smoking status, domiciliary oxygen therapy, lung function, symptom burden, and current medication use) to quantitatively predict 12-month risk of exacerbations and has been validated in real-world and clinical trial populations.^24,-28^ Through this intervention, the ACCEPT model will be integrated into electronic medical records at point-of-care and its clinical impact evaluated in a stepped-wedge cluster randomized trial (the IMPACT study)^23^. The physician component of the ADI uses the ACCEPT algorithm to improve appropriate prescribing by suggesting the recommended treatment based on clinical guidelines, while the patient component seeks to improve adherence to treatment by addressing beliefs about the prescribed medication and providing additional resources relevant to self-management of COPD.^10,11,29^ This paper describes the development process for the ADI, the core intervention in the ongoing IMPACT trial.^23^

## 2. Methods

### 2.1 Overall Approach & Methods

This study developed an integrated physician and patient decision tool for use in the IMPACT trial.^23^ Tool development was grounded in the *Knowledge To Action* (KTA) framework.^30^ The KTA framework is a widely recognized process model for knowledge translation research. It consists of two core components: knowledge creation and the action cycle. In this study, knowledge creation began with the development of the ACCEPT prediction model. We then aimed to refine this knowledge further and enable its use in clinical practice through the development of a knowledge tool: the ADI. In alignment with the KTA cycle, the tool’s development began following identification of a problem (i.e., inconsistent implementation of guidelines, and suboptimal treatment adherence) through a comprehensive review of the literature and existing systematic reviews (e.g., Sehl et al., 2017), and consultations with both clinicians and patients.^31^ This paper describes the following phases of the action cycle: adapting knowledge to the local context; assessing barriers and facilitators to knowledge use through user interviews and literature review; and selecting and tailoring the intervention. For the latter, we applied the Theory and Techniques Tool— an interactive evidence-based resource that links specific behaviour change techniques with mechanisms of action.^32–34^ This Tool guided intervention design by mapping barriers and facilitators to known behavioural change techniques. The stepped-wedge trial of the ADI will address implementation, knowledge use monitoring, and outcome evaluation.

Complementing the KTA framework, tool development followed an integrated knowledge translation approach whereby end-users drove the content and design of all materials. Research shows that engaging users in tool development can improve uptake.^35^ Thus, we began by developing a team of decision scientists and target tool users (i.e., clinicians and patient partners). ADI tool development followed an iterative, user-centered design approach^36^ and took place in three stages: 1) Information gathering and intervention selection; 2) Development of prototype tools; and 3) Assessment and revision of prototype tools.

This study was reviewed and approved by the University of British Columbia Behavioural Ethics Board (H21-02348).

### 2.2. Stage 1: Information Gathering and Intervention Selection

Semi-structured interviews were conducted with potential tool users (i.e., people with COPD and pulmonologists) to understand how pulmonologists and individuals with COPD make decisions about treatment, which considerations they prioritize in decision making (e.g., symptom reduction; treatment frequency), and barriers and facilitators to treatment uptake and adherence. Participants were recruited from the respiratory clinics at two tertiary care teaching hospitals, St. Paul’s Hospital and Vancouver General Hospital in British Columbia, through word-of-mouth and via organizations focused on lung health (e.g., Legacy for Airway Health). Recruitment aimed to maximize variation in experience. Based on the literature^37^ and feedback from patient partners on the research team, particular attention for recruitment of individuals with COPD was paid to diversity in age, gender identity, and time since diagnosis. Eligibility criteria, confirmed by a screening questionnaire, included: of at least 18 years of age; could communicate in English; and had a clinical diagnosis of COPD. Healthcare providers were eligible to participate if they were a pulmonologist providing care to individuals with COPD in Canada. Recruitment continued until saturation – that is, until interviews no longer revealed new insights.^38^

Due to pandemic restrictions, interviews were conducted remotely via telephone or Zoom, per participant preference. The interview guides were developed collaboratively by team members with expertise in decision science, knowledge mobilization, and smoking cessation in conjunction with those with lived experience of COPD. Two interviewers, both with previous experience in semi-structured interviewing in clinical research, completed all interviews. Member-checking was used throughout the interviews to ensure alignment between participants’ understanding of their own experiences and the qualitative data collected.^39,40^ The interviewers met regularly to address any issues that arose, identify commonalities and emerging findings to probe further, and to revise the interview guide as needed.

Interviews were audio-recorded and then transcribed. Audio files were kept until transcription was completed and then deleted. Transcripts were de-identified to protect participant confidentiality. All files were stored in accordance with the University of British Columbia’s Behavioural Ethics Board best practices. Memoing was conducted after each interview, with a focus on the most important considerations/topics to address, barriers to decision-making and action, and information preferences. Specifically, at the end of each interview, interviewers compiled a list of all barriers, enablers, and content areas identified as important by the interviewee. For both the provider and patient components of the tool, the list was built across successive interviews with intended tool users, with new content areas added as they arose. In line with a rapid qualitative approach,^41^ these memos served as the primary basis for analysis. Based on extensive evidence showing a relationship between gender and COPD symptoms and trajectory, analysis considered a sex and gender lens.^42–44^ Findings were shared with patient partners and the knowledge mobilization team throughout the interview process to assess for possible issues related to transferability of findings and to determine if revisions were required to the interview guide.^45^

Then, identified barriers/enablers were complemented by a search of the literature, and relevant domains were then mapped to possible behaviour change techniques through the Theory and Techniques Tool. Next, based on the behaviour change techniques that had the highest levels of evidence of effectiveness, patient partners and pulmonologists on the study team sought to identify a feasible intervention strategy encompassing those techniques. In determining intervention content, the complete list of all content areas identified in this phase, including illustrative quotes from interviews, was discussed and reviewed with patient partners and pulmonologists on the study team to determine which content should be included in the patient and physician components of the ADI.

### 2.3 Stage 2: Development of Prototype Tools

We developed prototypes of each ADI component (a decision support tool for physicians and an individualized decision handout for patients). Development of the patient component followed established International Patient Decision Aid Standards where applicable.^46,47^ As described above, tool content areas were based on the preferences and decisional needs identified in semi-structured interviews with target tool users. These content areas were reviewed by patient partners and pulmonologists on the study team for accuracy, relevance and completeness before prototypes were developed. Subject area experts guided development of tool content. Prototype tools were reviewed by intended tool users and subject area experts on the study team before feedback was sought from external users.

### 2.4. Stage 3: Assessment & Revision of Prototype Tools

Feedback was sought from intended tool users to refine the prototype tools and ensure they met user needs. Feedback was elicited in two different ways: pulmonologists completed an online survey; and individuals with COPD completed cognitive interviews (see below).^48^

#### 2.4.1. Online Survey for Pulmonologists

All pulmonologists at the two trial sites (n = 36) were invited by e-mail to complete a brief online survey to provide input on the prototype physician tool. Feedback was elicited via an anonymous, online survey rather than individual interviews to provide each pulmonologist with the opportunity to freely contribute to the development of the physician component of the ADI.

The online survey included a short video demonstration of the prototype tool, followed by brief demographic questions, the relevant items from the Ottawa Acceptability Scale (practitioner version),^49^ and open-ended questions assessing participant concerns about, and desired changes to, the tool.

#### 2.4.2. Cognitive Interviews with People with COPD

Participants were recruited using the same methods described above (Section 2.2) to complete one 45-to-60-minute cognitive interview. Individuals were eligible if they: were 18 years of age or older; could read and speak English; and had a diagnosis of COPD. Interviews were conducted in-person or via Zoom, per participant preference.

Two interviewers conducted all interviews. The cognitive interviews were semi-structured and recorded.^48^ As before, audio recordings were stored in compliance with the University of British Columbia Behavioural Ethics Boards’ best practices. In addition, the interviewers took notes during each interview and recorded thoughts and impressions before and after each interview. These notes served as the primary data source; audio recordings were consulted as needed to clarify notes. The interviewers held discussions after each interview to determine what revisions, if any, might be appropriate, and identify areas to probe further in subsequent interviews.

During the interviews, the prototype patient informational handout was presented to participants. Participants were instructed to read the handout and share their thoughts aloud. As needed during the interview, the interviewer would prompt the participant on specific aspects of the handout that were identified as important by the research team, pulmonologists, or previous participants. Interviews were designed to solicit feedback on layout, understandability of the figures and language, and appropriateness of the content, and to identify any missing elements. When applicable, participants were presented with previous versions of the handout to assess the acceptability of changes that had been made in response to previous participants’ suggestions.

## 3. Results

### 3.1. Stage 1: Information Gathering

In total, 5 pulmonologists and 17 people with COPD completed semi-structured interviews on treatment decision-making, barriers and enablers, and informational needs. Pulmonologists identified primarily as women (n = 4) and reported an average of 9 years of clinical practice (range: 2 years to 15 years). Participants with COPD ranged in age from 40 to 81 years; reported an average of 12 years since diagnosis (range: 1 year to 31 years); and 47% (n = 8) identified as women (Table 1).

**Table 1:** Demographics of Participants with COPD.

|  | **Stage 1: Information Gathering**<br>n = 17 | **Stage 3: Assessment and Revision of Prototype Tools**<br>n = 4 |
|---|---|---|
| **Gender identity % (n)** |  |  |
| Man | 53% (9) | 43% (3) |
| Woman | 47% (8) | 57% (4) |
| Prefer to self-describe | 0% (0) | 0% (0) |
| **Mean age (range)** | 67 years<br>(50-81 years) | 68 years<br>(55-76 years) |
| **Mean time since diagnosis (range)** | 12 years<br>(1-31 years) | 19 years<br>(7-35 years) |
| **Born in Canada % (n)** |  |  |
| Yes | 100% (17) | 100% (7) |
| **Comorbidities % (n)**<sup>\*</sup> |  |  |
| High blood pressure | 41% (7) | 14% (1) |
| Diabetes | 6% (1) | 14% (1) |
| Anxiety | 53% (9) | 29% (2) |
| Depression | 24% (4) | 43% (3) |
| Insomnia | 6% (1) | 14% (1) |
| Anaemia | 0% (0) | 0% (0) |
| **Annual household income % (n)** |  |  |
| Less than \$20,000 |  | 29% (2) |
| \$20,000 to \$39,999 |  | 29% (2) |
| \$40,000 to \$59,999 |  | 0% (0) |
| \$60,000 to \$79,999 |  | 14% (1) |
| \$80,000 to \$99,999 |  | 0% (0) |
| More than \$100,000 |  | 14% (1) |
| Prefer not to answer |  | 14% (1) |
| **Highest level of education completed % (n)** |  |  |
| Primary school | 6% (1) | 0% (0) |
| High school | 53% (9) | 43% (3) |
| College diploma or technical/trade school | 18% (3) | 43% (3) |
| University – undergraduate | 24% (4) | 14% (1) |
| University – graduate | 0% (0) | 0% (0) |
| Professional qualification | 0% (0) | 0% (0) |
| Other | 0% (0) | 0% (0) |
| Prefer not to answer | 0% (0) | 0% (0) |
| **Preferences for shared decision making % (n)** |  |  |
| I prefer to make the decision about which treatment I receive | 0% (0) | 14% (1) |
| I prefer to make the final decision about my treatment after seriously considering my doctor’s opinion | 18% (3) | 14% (1) |
| I prefer that my doctor and I share responsibility for deciding which treatment is best for me | 59% (10) | 57% (4) |
| I prefer that my doctor makes the final decision about which treatment will be used but seriously considers my opinions | 24% (4) | 14% (1) |
| I prefer to leave all decisions regarding treatment to my doctor | 0% (0) | 0% (0) |

<sup>\*</sup>responses missing for two Stage 3 participants

Several barriers and enablers were identified in the interviews. Pulmonologists reported difficulty identifying exacerbation history – the primary clinical measure used to assess and categorize patient risk in clinical guidelines. When asked about the ACCEPT model, they expressed a desire to see the model inputs, such as forced expiratory volume in one second (FEV1), as well as the numeric (vs. categorical) ACCEPT risk prediction. Pulmonologists emphasized that the tool should not add more work to the clinical encounter, especially given time constraints. Patients identified gaps in knowledge surrounding medication side effects and expected benefits, general understanding of COPD, inhaler technique, and common medical terms. They expressed a need for resources and additional supports surrounding exercise, including pulmonary rehabilitation; peer support; and smoking cessation. No apparent differences emerged in barriers or preferred supports by gender. Patient partners on the research team further suggested an additional module providing individualized information to help each patient understand their current lung function and exacerbation risk. Pulmonologists wanted the option to modify the content of the patient handout based on their discussion with each patient.

Broadly, the study team determined that the barriers reported by physicians and patients could be addressed by techniques addressing the knowledge, skill, behavioural cueing, and general attitudes/beliefs mechanisms of action in the Theory and Techniques Tool. Thus, relevant and evidence-based behaviour change techniques were selected to guide design of the physician and patient tools. For physicians, these included: information about health consequences; prompts and cues; and conserving mental resources. Relevant behaviour change techniques for patients included: information about health consequences; salience of consequences; comparative imagining of future outcomes; social support; instructions on how to perform behaviour; and behavioural practice/rehearsal. Based on review of these techniques and clinical operations, a computerized decision support intervention was selected as the optimal format for physicians. For patients, a decision aid was considered but rejected due to the limited time allotted for appointments. Instead, it was determined that an individualized paper informational handout was the optimal format to balance patient and pulmonologist needs with clinic operations.

### 3.2. Stage 2: Development of Prototype Tools

The initial prototype pulmonologist component of the tool was designed as an electronic decision support tool that collected data on the core ACCEPT input variables of FEV1, age, sex, height, weight, and smoking status directly from the electronic health record (EHR), where possible. The tool then provided an individualized risk prediction for each patient. Additionally, based on medication history and reported exacerbations, the tool showed where the patient was in the Canadian Thoracic Society (CTS) algorithm, and highlighted the medication recommended by the CTS COPD guidelines. In particular, CTS recommendations are based on the binary classification into low and high risk for future exacerbations were mapped to the ACCEPT numerical prediction using a risk threshold. To objectively derive this threshold, the Evaluation of COPD Longitudinally to Identify Predictive Surrogate Endpoints (ECLIPSE) cohort was analyzed to find the threshold on predicted risk that results in the same proportion of high-and low-risk individuals as the CTS’ exacerbation-based definition.^26,50^ Treatment used in prior exacerbation(s) and exacerbation history were a priori designed to require direct physician input because both fields require clinical judgment and be reliably retrieved from electronic charts.

Feedback from the study team resulted in several changes to the prototype: the number of steps to complete the tool were reduced; the phrasing of medication de-escalation recommendations was changed to more accurately reflect the CTS guidelines; the layout of the recommendation was moved to appear below a diagram of the CTS algorithm; and a mandatory prompt was added to require pulmonologists to confirm their decision if they prescribed a medication not recommended by the CTS guideline. Figure 1 shows the prototype pulmonologist component of the tool.

**Figure 1:**
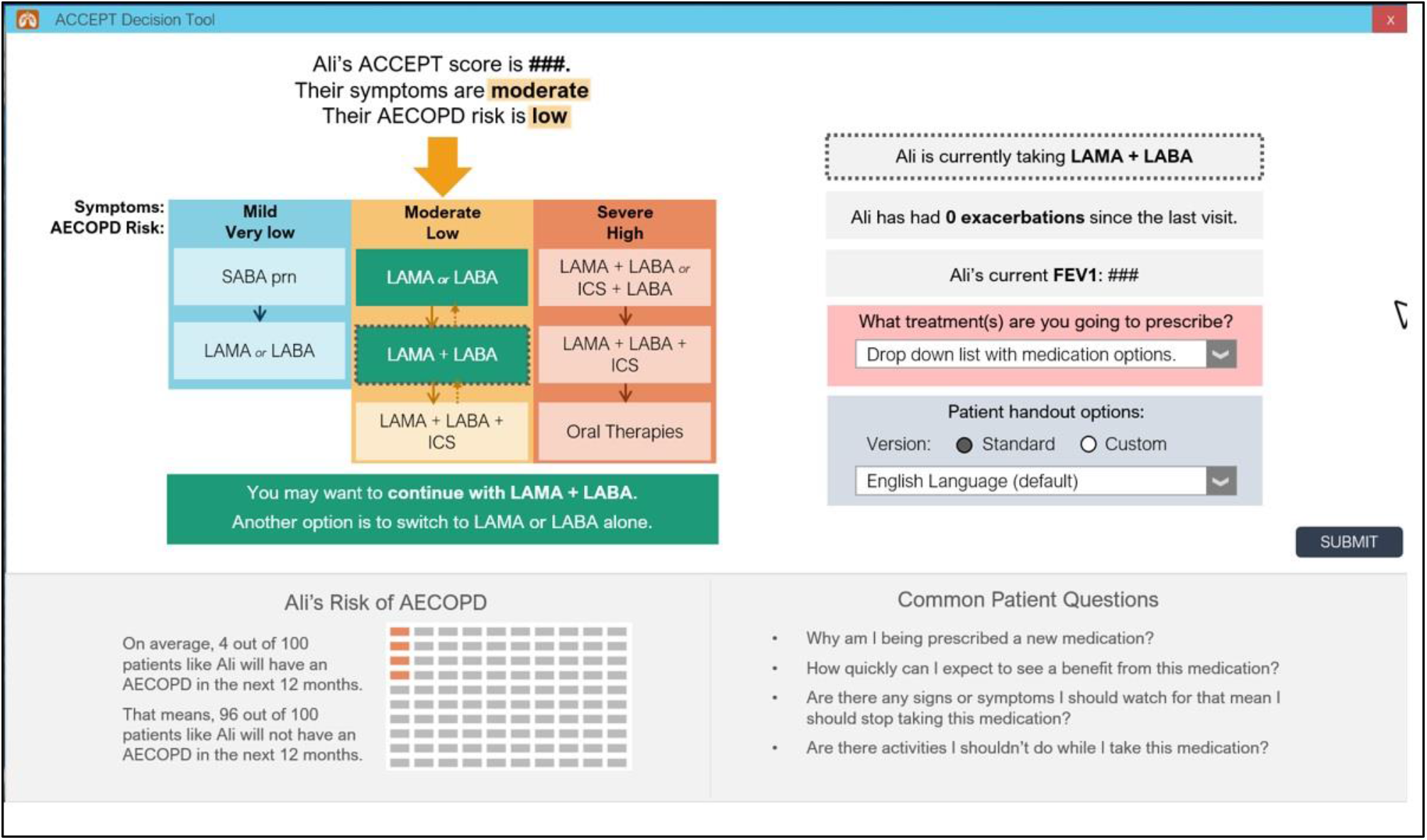
Main Screen of Physician Tool Showing Patient Place in Clinical Guidelines

The initial prototype of the patient component of the ADI was designed as a 2-page (4 sides) modular handout to align with the information and decision making that had been discussed with the previous physician component of the tool. It provided individualized information to each patient based on their most recent spirometry results, their ACCEPT risk prediction, their prescribed maintenance medication, and guidance related to COPD self-management. There were eight modules included—one for each content area identified in Stage 1 as well as a module called “My COPD” that provided individualized information on current lung function and risk of a severe exacerbation without treatment.

The handout was designed with a standard layout and module order. Pulmonologists could customize which modules were included using the pulmonologist component of the tool. The smoking cessation module was modelled on a validated infographic utilizing a lung function decline prediction calculation (a personalized biomedical risk assessment) designed to be used to support smoking cessation conversations between physicians and patients with COPD.^51^ To make the infographic more accessible, chunking was used:^52^ the validated infographic was divided in two with the introductory part of the infographic presented in the first module (My COPD) to explain recent spirometry results; and then the full infographic presented in the smoking cessation module. Feedback from subject matter experts led to revisions in the smoking cessation module to explicitly acknowledge gender differences in the effectiveness of smoking cessation tools. Following feedback from patient partners and knowledge translation experts, the order of the modules in the standard layout was revised; some module content was reframed to be more positive; and the title font for each module was changed to improve legibility.

### 3.3. Stage 3: Assessment & Revision of Prototype Tools

#### 3.3.1. Online Survey for Pulmonologists

Seventeen of the 36 invited pulmonologists watched the demonstration video and completed the online survey (response rate: 47%). In general pulmonologists viewed the physician component as acceptable, with all or almost all agreeing that the physician tool was: easy to understand (94% of respondents); compatible with the way things should be done (88%); and complementary to their usual approach (82%). Most pulmonologists were either unsure if the tool would save time (35%) or thought it would not save time (39%; see Table 2 for detailed results). This concern about the time required to complete the tool was apparent in pulmonologist responses to open-ended questions: several suggested changes to the tool focused on decreasing pop-up boxes and data-entry requirements, and making sure the tool was well integrated into the clinical workflow. In addition to concerns about time requirements, pulmonologists expressed concern that the tool was too simple and would not match the complexity of some patients, and that the EHR data collected would not be reliable. Lastly, pulmonologists requested several changes including a way to integrate the tool with clinical notes, and decreased data entry requirements and mouse clicks.

**Table 2:** Acceptability of Prototype Pulmonologist Tool (n = 17 pulmonologists)

|  | **Strongly Disagree** |  |  |  | **Strongly Agree** |
|---|---|---|---|---|---|
|  | **1** | **2** | **3** | **4** | **5** |
|  | % (n) | % (n) | % (n) | % (n) | % (n) |
|---|---|---|---|---|---|
| This tool will be easy for me to use | 6% (1) | 0 (0) | 24% (4) | 29% (5) | 41% (7) |
| This tool is easy for me to understand | 0 (0) | 6% (1) | 0 (0) | 47% (8) | 47% (8) |
| This tool is compatible with the way I think things should be done | 0 (0) | 0 (0) | 12% (2) | 47% (8) | 41% (7) |
| Using this tool with save time | 0 (0) | 29% (5) | 35% (6) | 18% (3) | 18% (3) |
| This tool complements my usual approach | 0 (0) | 0 (0) | 18% (3) | 59% (10) | 24% (4) |
| Using this tool does not involve making major changes to the way I usually do things | 0 (0) | 12% (2) | 6% (1) | 59% (10) | 24% (4) |
| This is a high probability that using this tool may result in more benefit than harm* | 0 (0) | 0 (0) | 25% (4) | 44% (7) | 31% (5) |

<sup>\*</sup>16 participants responded to this question

The physician component of the ADI was revised based on this feedback so that there were fewer mouse clicks and required data fields were clearly labelled and organized at the top of the screen to reduce the amount of effort needed to complete the tool. Further, although it was not possible to integrate it with EHR clinical notes, a feature was added so that the tool automatically generated a clinical note at the end of the encounter based on earlier pulmonologist inputs which could then be copied and pasted into the EHR (see supplementary for a video overview)

#### 3.3.2. Cognitive Interviews with People with COPD

Seven people with COPD completed cognitive interviews to elicit feedback on the prototype patient tool. Participants were between 55 and 76 years of age and first received their COPD diagnosis between 7 and 35 years prior. Four identified as women (Table 1). Participants provided feedback on all aspects of the prototype patient component of the ADI. Feedback was focused primarily on the format and phrasing of content rather than the content itself. For example, participants reported a desire for more visual aids and less text. As a result, text descriptions of COPD symptoms and tips for proper inhaler use were replaced with graphics.

Participants suggested changes to phrasing to make the handout more inclusive by adding language acknowledging that exercise can look different for each person; and clearer by specifying that the medication information pertained only to maintenance medications prescribed at the present appointment. Although at least one physician in earlier interviews raised concerns that including content on smoking cessation could be alienating, interviewed participants did not share this concern. Instead, they reported that including information on smoking cessation would be helpful and could make it easier for patients to start conversations about quitting smoking with healthcare providers.

Based on participant feedback, significant revisions were made to the smoking cessation infographic included in the smoking cessation module. Participants and physician team members expressed concern that the infographic contained too much information to be accessible to patients reviewing the infographic on their own. A revised graphic was designed that combined the information provided in the initial smoking cessation graphic, with the formatting of another validated graphic designed to explain lung function results to patients.^53^ Chunking was retained: half of the graphic was first presented in the *My COPD* module explaining spirometry results; and the full graphic was presented afterwards in the smoking cessation module. The revised smoking cessation graphic is shown in Figure 2. A video overview of the revised tool is provided in the supplementary material.

**Figure 2:**
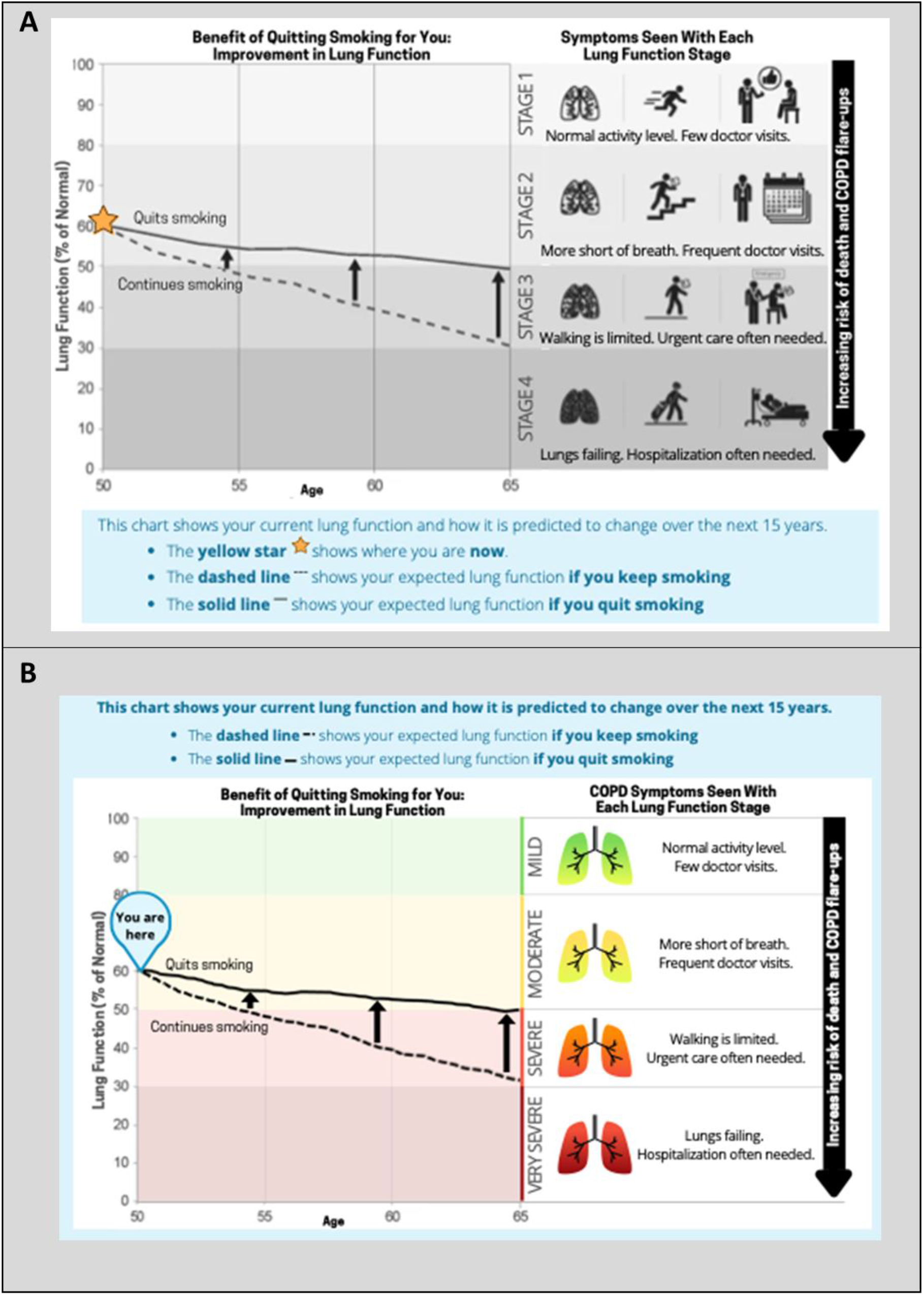
Changes to Smoking Cessation Infographic Following Cognitive Interview

## 4. Discussion

This paper reports the development of an integrated decision tool with two components: a computer-based decision intervention to support physicians in providing guideline adherent care, and an individualized educational handout to support patients in medication adherence and self-management. The intervention was designed in accordance with the KTA framework. Patient and provider barrier and enabler assessment, complemented by literature review, allowed us to identify relevant behaviour change techniques, which then led to the selection of the appropriate intervention type for each user type. Tool development itself also followed a rigorous process. Content and format of the tool were tailored to address the most relevant barriers, including physician knowledge and time limitations preventing accurate risk categorization and patient misconceptions regarding medications and a lack of knowledge about their benefits. Most importantly, to improve risk categorization, the tool incorporates a clinical prediction model instead of classification based on exacerbation history alone; the latter is shown to be at high risk of causing clinical harm.^54^ To support positive patient beliefs about medication taking, the tool includes accessible information about the risks and benefits of the maintenance medication being prescribed, individualized risk predictions, how to use the medication effectively, and how to monitor response over time. The development of the patient component followed international guidelines (where applicable),^46,47^ and both components were designed according to user-centered design principles.^36,55^ Development relied on evidence-based strategies including: team-based design which included end-users; observational field visits; interviews about user perspectives; quantitative surveys with potential users; iterative processes; and live prototyping (or cognitive interviewing).^35^

The patient component of the ADI aligns with previous studies of patient experiences with COPD reporting that the impact of COPD on quality of life is significant and can be underestimated.^56^ Patients interviewed in the information gathering stage of our study reported a desire for content that could support people with COPD to improve their quality of life by maintaining or building physical wellness through healthy behaviours such as increasing exercise and reducing smoking. As a result, the patient component of the ADI included resources on pulmonary rehabilitation and smoking cessation. As a paper tool, the patient component of the ADI diverges from newer trends in digitization of patient tools. While digital tools allow a greater level of personalization which can increase tool uptake,^57^ we developed the patient component of the ADI as a paper tool based on feedback from clinic staff, pulmonologists and patients. This feedback identified a digital tool as a potential barrier to uptake due to uneven access to equipment (i.e., smart technology) and technological literacy in the target patient population. This is in-keeping with prior studies showing decreased uptake of electronic questionnaires, particularly in elderly populations (comprising many patients with COPD).^58^ We attempted to balance the need for the paper-based tool with the personalization advantages offered by a digital tool by leveraging patients’ recent lung function test results to generate personalized risk predictions that were embedded in the handout. Additionally, the modular structure of the tool allows pulmonologists to tailor the tool content to each patient by adding or removing modules that are more or less relevant to each individual patient.

Existing evidence shows that despite the increasing spread of EHRs and quickly advancing technology, the uptake of clinical decision-support tools by physicians is inconsistent.^59^ In order to overcome barriers to adoption like alert fatigue,^60^ clinical decision support tools need to be easy to use and offer real value to intended tool users.^61^ We aimed to maximize the value proposition of the physician component of the ADI through user-centered design and by creating a tool that directly addressed target end-users needs. Through iterative testing, we adapted the tool to better fit into the physician workflow. However, more thorough integration, such as native EHR-integration, may have further supported adoption and uptake. Future work should address how best to optimize integration of decision support tools like the ADI into various EHRs, made more feasible through increasingly common open application programming interfaces (i.e., APIs) leveraging the widespread Fast Healthcare Interoperability Resources standard. Almost all pulmonologists surveyed indicated that the tool was easy to understand and complemented their usual approach. However, one of the major challenges to the uptake of clinical decision support tools in chronic diseases, specifically, is a mismatch between patient complexity and tool scope.^61^ That is, although many patients have multimorbidity, clinical decision support tools tend to focus on a single condition, potentially restricting their applicability in everyday clinical practice. Pulmonologists participating in this study shared this concern about the ADI, which may ultimately limit its uptake and impact. Future work will explore potential adaptations to the ADI to better meet the needs of patients with multimorbidity. This might include using EHR records to identify patients with clinically important multimorbidity and tailoring the tool to signal to the physician that specific consideration is required. This adaptation is of particular importance given the potential that decision tools like the ADI could oversimplify clinical decision-making to the detriment of the patient. These findings are consistent with studies exploring barriers to implementation of computerized decision support that emphasize the importance of fit with workflow, usefulness in practice, technical dependencies and design, and the contextual fit of the tool with the user’s role or clinical setting.^62^

### 4.1. Limitations

This development process lays the foundation for the successful implementation of the integrated tool, however, there are limitations. First, the ACCEPT model was developed based on different populations than those included in the planned IMPACT trial. As such, it is possible that the model may not be well-suited to the clinical trial population. To assess this empirically, the IMPACT study will compare the predicted risk of exacerbation from the ACCEPT model and the observed exacerbation risk among the IMPACT trial population. Second, the ADI was developed within a Western Canadian context; the majority of people with COPD and pulmonologists who contributed to the tool development were residing in British Columbia. To address this, region-specific content, such as the smoking cessation resources, can be easily adjusted to reflect relevant resources for other jurisdictions. Relatedly, participation in this study was limited to English-speaking individuals, despite the linguistic diversity present in British Columbia and evidence that non-anglophones may benefit more from a decision support tool. Future work should explore adaptations to meet the needs of non-anglophone populations. Furthermore, the software component of the tool is intentionally detached from the EHR to allow implementation in different clinical settings, such as primary care where COPD is often treated. During development, potential adoption by a wide variety of EHR systems was considered. As a result, the ADI’s core is vendor agnostic; it uses a flexible pipeline based on optical character recognition for reading text data from the screen, enabling direct data capture regardless of EHR system. However, full EHR integration of this tool should be explored as this may improve uptake by reducing physician burden. Additionally, all pulmonologist participants worked at large research hospitals. Although the needs of non-academic pulmonologists may differ, the clinical guidelines and risk calculator upon which the pulmonologist component is based are universal. In addition, the ADIwas designed for use in specialty care and for more severe disease. As such the tool may not be appropriate for primary care, where the majority of patients with COPD are managed. Adapting the tool for primary care would require consultation with primary care clinicians as well as pharmacists who increasingly support outpatient management of COPD.^63,64^ Full adaptation would need to consider barriers and facilitators to implementation that are unique to the primary care context, such as the availability of input variables required for the ACCEPT model. Ideally such a model could be integrated into a point-of-care computerized decision support system which could facilitate assessment of symptom burden and exacerbation risk, offer specific drug name and dose suggestions corresponding to ACCEPT class change recommendations, and enable proven self-management/educational interventions such as pre-population of COPD action plans. Lastly, user-centered design does not guarantee the diffusion and uptake of clinical tools. Furthermore, participation in one stage of tool development was not an exclusion for participation in the subsequent stage, potentially decreasing the overall impact of the user-centered approach. That being said, challenges to uptake are multifold, and for clinical decision support tools in general include alert fatigue,^60^ time requirements, and poor physician buy-in. Despite evidence that much COPD prescribing is inappropriate, pulmonologist participants in Stages 1 and 3 raised concerns the tool may have low uptake among pulmonologists who perceive themselves to be delivering evidence-based pharmacotherapy already (a barrier this intervention does not address).

## 5. Conclusions

Guided by the KTA framework, this study employed a user-centered approach to develop a comprehensive decision intervention for COPD: the ADI. The ADI includes components tailored to the unique needs of pulmonologists and patients with COPD. The goal of the ADI is to improve health outcomes for patients with COPD by helping pulmonologist accurately assess patient risk for exacerbations to improve appropriate prescribing, and by supporting patients to adhere to prescribed medications. The specific interventions included in the ADI were evidence-based and informed by the Theory and Techniques Tool. Preliminary testing of the ADI with target end-users found that the tool was easy to understand and perceived as relevant. However, pulmonologists expressed concern about the reliability of the EHR data on which the tool relies. Future work using qualitative and quantitative methods will evaluate the impact of this integrated tool on physician prescribing, patient engagement in treatment decision making, and patient adherence, in a stepped-wedge trial.^23^

## List of Abbreviations

ACCEPT: Acute COPD Exacerbation Prediction Tool
ADI: Accept Decision Intervention
COPD: Chronic obstructive pulmonary disease
CTS: Canadian Thoracic Society
EHR: Electronic health record
FEV1: Forced expiratory volume in one second
KTA: Knowledge To Action

## 6. Declarations

### 6.1. Ethics approval and consent to participate

This study was reviewed and approved by the University of British Columbia Behavioural Ethics Board H21-02348. All participants provided informed consent prior to participation.

### 6.2. Consent for publication

Not applicable

### 6.3. Availability of data and materials

Qualitative datasets used in the study are not publicly available due to privacy considerations. Quantitative data are included in the present publication.

### 6.6. Acknowledgements

We wish to thank all the patients and respirologists that participated in this study.

### 6.7. Author Contributions

MS, DS and NB acquired funding for the project. NB, RM, MS, AH, SG, JB, LS, JJ and DS were involved in developing and conceptualising the research. PB was responsible for the project administration. RM and RS collected the data, which was analysed by RM under the supervision of NB and MS, with support from AH, JB, LS, SG, JJ and DS. MM developed the interface. RM wrote the initial manuscript and prepared the figures together with NB. All authors contributed to the article by means of critical suggestions and revisions, and have approved the final manuscript.

