## Supplementary material for "Development of Integrated Physician and Patient Tools to Improve Management of COPD: The ACCEPT Decision Intervention"

1. **Video Overview of ACCEPT Decision Intervention**

A video overview of the ACCEPT Decision Intervention is available at the following link: <https://ln5.sync.com/dl/d1b4dcb70/view/video/34948779900102?sync_id=0#x7zzbc97-jtmywvc8-eycrmqrx-aszzxuk9>
